# Improving Knowledge and Practices on Genital Chlamydia among Youths aged 15–24 Years Old in Bamako: Evaluation of a 5-Day Hybrid Workshop

**DOI:** 10.64898/2026.03.09.26347950

**Authors:** Modibo Sangaré, Bekaye Coulibaly, Kassoum Alou Ndiaye, Fatoumata Doumbia, Drissa Konate, Karim Traore, Seidina A Diakite, Dramane Sogodogo, Kletigui Casimir Dembélé, Mody Cissé, Samba Diarra, Romain Dena, Bourama Keita, Abdrahamane Anne, Emile Badiel, Daouda S Niare, Mariam Toure, Moussa Niare, Adama Ouedraogo, Désiré Tassembedo, Mamady Traore, Madina Konaté, Mamadou Diop, Daouda Fomba, Oumar Sidibé, Modibo Kouyate, Aissatou Soumana Billo, Privat Agniwo, Christelle Fatondji, Ichiaka Moumine Koné, Hama Diallo, Boulaye Sanogo, Ousmane Traore, Ousmane Maiga, Abdrahamane Anne, Housseini Dolo, Lalla Fatouma Traore, Kassoum Kayentao, Souleymane Dama, Sekou Bah, Boureima Guindo, Sory Ibrahim Diawara, Fatoumata Traore, Cheick AT Traore, Kalifa Keita, Fumiko Shibuya, Jun Kobayashi, Mahamadou Diakite

## Abstract

**Introduction:** Genital chlamydia is one of the most common sexually transmitted infections (STIs) among young people aged 15–24 years old and may lead to serious complications in women, men, and newborns. This study aimed to evaluate the effectiveness of a five-day interactive hybrid educational workshop on knowledge, attitudes, and practices (KAP) related to genital chlamydia in Bamako, Mali.

**Materiel and Methods:** A descriptive pre-test/post-test study was conducted in 2025 among 174 young participants aged 15–24 years old. Data were collected using a standardized questionnaire assessing KAP related to genital chlamydia and STIs. Proportions before and after the intervention were compared using the chi-square (χ²) test or Fisher’s exact test, with a significance level set at α = 0.05.

**Results:** Overall knowledge of STIs showed a slight improvement after the intervention, with the “poor knowledge” category disappearing at post-test. The proportion of participants able to identify at least three modes of transmission of genital chlamydia increased from 41.7% to 50.0%. Improvements were also observed in the identification of symptoms and complications, with a statistically significant increase in knowledge of male complications (p = 0.045 for at least one complication; p = 0.003 for at least two complications). Information sharing about genital chlamydia with partners remained limited (8.3% vs. 7.4%), whereas discussion of STI status between partners increased (62.5% to 72.2%). Preventive practices improved modestly, including consistent condom use (62.5% to 66.7%), prompt health-care seeking (95.0% to 96.3%), and acceptability of on-demand STI screening (70.8% to 83.3%).

**Conclusion:** The hybrid educational workshop contributed to improved knowledge and several positive attitudes toward genital chlamydia, particularly regarding communication about STI status and acceptability of screening. However, persistent stigma and limited information sharing with sexual partners underscore the need for sustained and repeated educational interventions targeting young people for chlamydia prevention and control.

## 1. INTRODUCTION

Adolescent sexuality in Africa has historically been framed by strict traditional norms, centered on marriage and procreation, and marked by strong taboos regarding sex outside marriage. Colonization and the establishment of monotheistic religions have profoundly modified these norms, reinforcing the discourse of guilt around sexuality, particularly female sexuality (Dialmy, 2005). More recently, globalization and the rise of digital technologies are exposing young people to new relational and sexual models, often inspired by Western cultures, creating tensions between traditional norms and contemporary practices (Chambon, 2022).

These socio-cultural changes are accompanied by a weakening of social control over the onset of sexual life and a lengthening of the period of premarital sex (Delaunay V & Guillaume A, 2007). At the same time, parent-child communication on sexuality remains limited, with issues about sexuality and sexual health remaining largely taboo within African families (Hien et al., 2012). However, several studies have shown that an open parental dialogue about sexual risks is associated with delayed first sexual intercourse and a reduction in risky behaviours (Hutchinson et al., 2003).

Due to their physiological and behavioural development, adolescents are particularly vulnerable to risky sexual practices, exposing them to a high prevalence of STIs, including chlamydia. Caused by *Chlamydia trachomatis*, this infection is the most common bacterial STI in the world, with more than 105 million new cases estimated each year (Rowley et al., 2016; Duval et al., 2021) and 128.5 million new infections among adults aged 15 to 49 years old in 2020. In Europe, the incidence continues to increase, reaching 70.4 cases per 100,000 population in 2023 (ECDC, 2023).

In Africa, genital chlamydia remains largely underdiagnosed due to its often asymptomatic nature and the limitations of epidemiological surveillance systems (WHO, 2025a). Data are particularly scarce in North Africa and heterogeneous in sub-Saharan Africa, highlighting the need to strengthen screening and harmonize data collection methods, particularly in West Africa (Omori et al., 2025). In Mali, the available data mainly concern populations at intermediate risk between 2000 and 2009, leaving a significant lack of recent information on the general population and young people (Dembélé OD, 2012).

Genital chlamydia is most often asymptomatic, but can cause severe complications such as pelvic inflammatory disease, infertility, increased risk of HIV infection, and adverse pregnancy outcomes (WHO, 2025a; Institut Pasteur, 2024). Its prevention is based on the ABCDE strategy recommended by the WHO, including abstinence, fidelity, correct condom use, early detection and sex education (WHO, 2024a; WHO, 2025b). However, in resource-limited settings, diagnostic capacity remains insufficient, as nucleic acid amplification tests are not always available, which limits the effectiveness of management (Dalle Nogare, 2014).

In the face of these shortcomings, it is essential to strengthen prevention, improve knowledge and promote safer sexual behaviours among young people, while developing diagnostic and therapeutic capacities in Mali and Burkina Faso (Bocoum et al., 2017; Tovo et al., 2021). The objective of this study was to evaluate the effectiveness of hybrid educational workshops in terms of knowledge acquisition, positive change in attitudes and promotion of good practices among young people aged 15-24 years old in Bamako regarding STIs, in particular genital chlamydia.

## 2. MATERIAL AND METHODS

### 2.1. Study Design and Period

We conducted a descriptive study from June to December 2025. Data collection took place from August 16 to September 13, 2025.

### 2.2. Workshop Participant Recruitment

### 2.2.1. Eligibility Criteria

Applicants were eligible if they met the following criteria:

• **Age**
○ 18–24 years old for in-school and out-of-school youth; and/or
○ 25–49 years old for young health professionals.
• **Educational or Professional Status**
○ Formal enrollment in a secondary or tertiary institution; or
○ Recommendation by a recognized social or community structure for out-of-school youth.
• **Geographic Location**
○ Originating from or residing in Mali, Niger, Benin, or Burkina Faso at the time of application.
• **Language Proficiency**
○ Proficiency in French.
• **Technical Requirements**
○ Functional access to a smartphone, tablet, or computer with a stable internet connection for online participation.
• **Availability and Commitment**
○ Commitment to attend the entire workshop, either in person or remotely.

In addition, applicants were required to demonstrate a clear interest in sexual and reproductive health and STIs, particularly genital chlamydia, and to provide informed consent for voluntary participation and data collection.

#### 2.2.2. Selection Process

Applications that were incomplete or did not meet the eligibility criteria were excluded. Final participant selection aimed to ensure:

○ Balanced country representation, in accordance with the study protocol (i.e., ten participants from Mali and five participants from each of the other countries per face-to-face and online workshop);
○ Gender balance, with positive discrimination in favor of women and persons living with disabilities;
○ Diversity of academic and professional profiles; and
○ Inclusion of participants with strong potential for cascading dissemination of acquired knowledge and skills (K&S) within their communities and institutions.

### 2.3. Intervention Design

The study evaluated the effectiveness of three (3) hybrid educational workshops (face-to-face and online) conducted in August - September 2025. These workshops aimed to strengthen the knowledge, attitudes, and practices (KAP) of youths aged 15–24 years old regarding STIs, with a particular focus on genital chlamydia.

Workshop implementation was led by the Malian Network for Health, Education, Research and Development (MNHERD) in collaboration with the project investigators.

### 2.4. Pedagogical Approach

The workshops employed a participatory pedagogical approach emphasizing small-group work through small group facilitation. Face-to-face participants were organized into groups of five (5) to eight (8) individuals each, while online participants were assigned to virtual breakout rooms, each supervised by at least one co-facilitator.

For each activity, specific roles (a facilitator, reporter, timekeeper, observer, and a listener) were assigned to promote active engagement and accountability. Daily individual and group assignments focused on the reading and synthesis of the nine (9) chapters of a developed educational 280-page brochure, which served as the main workshop material. This brochure was shared with participants at least one week prior to the start of the workshops.

### 2.5. Workshop Content and Schedule

Each workshop was conducted over five (5) consecutive days:

○ **Day 1**: Presentation of workshop objectives and development of six key skills, including active listening, condom negotiation, SMART goal formulation, STI coaching (GROW/OSCAR models), structured brainstorming, and effective note-taking.
○ **Day 2**: Role-playing, interactive discussions, and lectures on STIs and genital chlamydia.
○ **Day 3**: Group presentations on STIs, discussions on the WHO ABCDE approach, and thematic conferences (traditional medicine, dermatology, zoonoses, and sexuality).
○ **Day 4**: Supervised development of community action plans and conference-debates on traditional sexuality education and menstrual hygiene.
○ **Day 5**: Presentation and evaluation of community action plans - considered the main indicator of knowledge appropriation and positive attitudinal change - by a jury using predefined evaluation criteria for K&S cascading prize (up to US$200 per group).

We declined the workshop agenda into sessions following the basic guidelines for sexual and reproductive health training for adolescents and youths:

○ *Each session should be anchored in the participants’ context, encouraging them to reflect on and share their experiences, using these as learning resources*.
○ *Adopt a positive, non-judgmental approach to sexuality that acknowledges it as a natural part of life and emphasizes the importance of expressing it without shame or taboo for individual well-being*.
○ *Create a safe space that fosters open, honest dialogue, ensuring that privacy and confidentiality are understood and respected*.
○ *Involve young co-facilitators or guest speakers who represent the participants’ peer groups and the session topics*.
○ *Be prepared for emotionally challenging moments and designate someone capable of supporting participants and facilitators when needed, while also attending to your own well-being with breaks and self-care*.
○ *Plan carefully but remain flexible to respond to unexpected group dynamics, and allow time for participants to discuss personal issues while maintaining balance for rest*.
○ *Encourage participants to lead energizing activities, promote critical reflection to identify stigmatizing practices, and foster personal responsibility for learning by preparing for sessions and reviewing discussions*.
○ *Be pragmatic and responsive to evolving group needs, adjust the schedule when necessary, and above all, ensure the sessions are engaging and enjoyable for everyone* (Stackpool-Moore, L. & Singh, A, 2015; Le Projet AgirPF. 2015; Stangl AL, Grossman CI, 2013).

### 2.6. Workshop Effectiveness Evaluation

A pre-test/post-test evaluation was conducted among 174 workshop participants. Workshop effectiveness was assessed based on:

○ Quality of group outputs (presentations and community action plans);
○ Level of active participation in discussions; and
○ Participants’ ability to design and propose activities aimed at disseminating acquired knowledge and skills to young people aged 15–24 years within their communities.

### 2.7. Data Collection and Analysis

We collected data using a 31-item structured questionnaire administered online. The questionnaire link was shared with participants via email and WhatsApp.

Proportions observed before (pre-test) and after (post-test) the workshops were compared using the chi-square (χ²) test or Fisher’s exact test, as appropriate. Statistical significance was set at α = 0.05.

Responses to questions assessing general knowledge and attitudes toward STIs, including genital chlamydia, were numerically coded (**Table 1a**). Overall scores were then categorized into three levels reflecting knowledge and attitude status (**Table 1b**).

**Table 1a.** Numerical Coding for Scoring Responses to Questions on General Knowledge and Attitudes toward STIs, Including Genital Chlamydia.

| # | Question | Response Options | Numerical Code |
| --- | --- | --- | --- |
| 1 | Is chlamydia a serious disease? | Yes / No / I don't know | Yes = 2; No = 1; I don't know = 0 |
| 2 | Should everyone be screened? | Yes / No / I don't know | Yes = 2; No = 1; I don't know = 0 |
| 3 | Would you consult a health professional? | Yes, immediately /<br>Yes, with hesitation /<br>No | Yes, immediately = 2; Yes, with hesitation = 1; No = 0 |
| 4 | Feeling ashamed to talk about it with one's partner | Yes / No / I don't know | Yes = 0; No = 1; I don't know = 0 |
| 5 | Stigmatization of people living with STIs | Yes / No / I don't know | Yes = 0; No = 1; I don't know = 0 |
| 6 | Would your partner agree to be tested? | Yes / No / Maybe / I don't know | Yes = 2; Maybe = 1; No = 0; I don't know = 0 |

**Table 1b.**
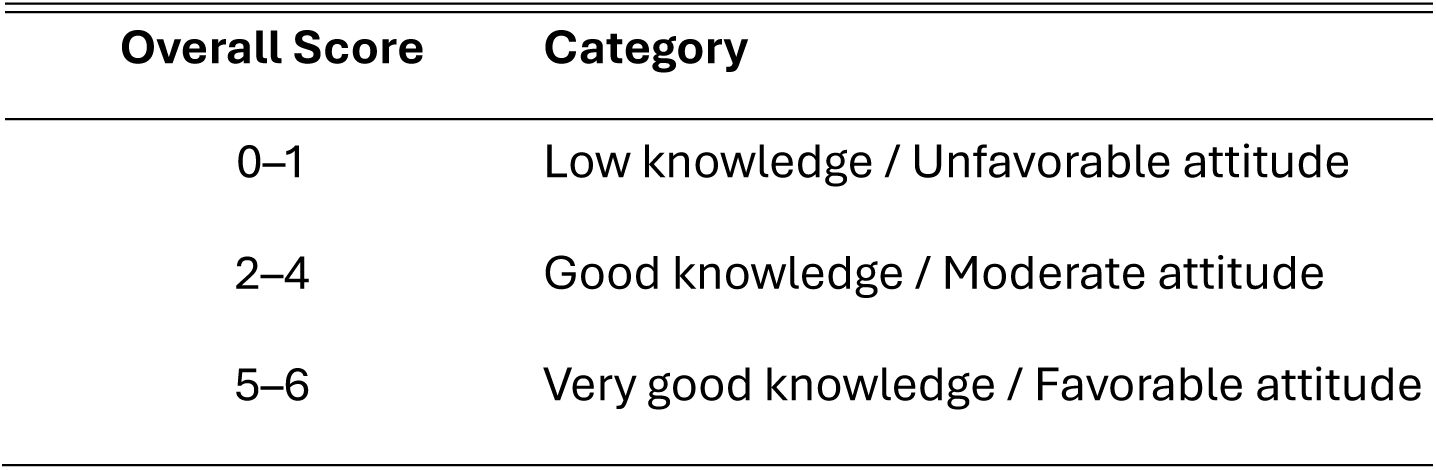
Categorization of Participants Based on Overall Scores for Knowledge and Attitudes toward STIs, Including Genital Chlamydia.

### 2.8. Ethical Considerations

We conducted this study in accordance with the ethical principles of the Declaration of Helsinki. The research protocol, data collection tools, and recruitment procedures were reviewed and approved by the Ethics Committee of the Hospital of Mali (Reference No. 2025-001) and the Ethics Committee for Health Research in Burkina Faso (Reference No. 2025-06-230).

Participation was entirely voluntary and conditional upon the provision of written or electronic informed consent for adults and parental consent for Malian minors prior to study inclusion. Participants were informed about the study objectives, procedures, potential benefits and risks, and their right to withdraw at any time without prejudice.

Participants received compensation for transportation, accommodation, and participation in accordance with rates approved by the Pfizer Faculty of Pharmacy (FAPH) Chlamydia Project Steering Committee. All data were anonymized, treated confidentially, and used exclusively for research purposes. Access to the data was restricted to two authorized senior research team members.

### 2.9. Some Key Acronym Definitions

**SMART (**Bahrami Z, Heidari A, Cranney J, 2022)

**S** – **S**pecific: clearly states *what* will be achieved, *by whom*, and *for whom*.

**M** – **M**easurable: includes indicators that allow progress and results to be quantified or assessed.

**A** – **A**chievable: realistic and attainable given available resources, time, and context.

**R** – **R**elevant: aligned with the identified problem and the overall purpose of the project or study.

**T** – **T**ime-bound: specifies a clear timeframe for achievement.

**GROW** (Whitmore J, 2017, Norman K, 2022)

**G** – **G**oal: Define a clear, specific, and achievable objective.

**R** – **R**eality: Explore the current situation, including challenges, resources, and constraints.

**O** – **O**ptions: Identify possible strategies or solutions to achieve the goal.

**W** – **W**ill (or **W**ay forward): Commit to concrete actions, responsibilities, and timelines.

**OSCAR** (Norman K, 2022; Downey M, 2014).

**O** – **O**utcome: Define the desired result or success criteria.

**S** – **S**ituation: Assess the current context and performance level.

**C** – **C**hoices (or Consequences): Explore available options and their potential consequences.

**A** – **A**ctions: Select and plan concrete actions to achieve the outcome.

**R** – **R**eview: Monitor progress, evaluate results, and adjust actions as needed.

**WHO ABCDE Approach** (WHO, 2024b)

**A** – **A**bstinence: Delaying the initiation of sexual activity or choosing not to engage in sexual intercourse, particularly among adolescents, as a means of preventing STIs.

**B** – **B**e faithful (or mutual fidelity): Maintaining a mutually faithful sexual relationship with an uninfected partner to reduce the risk of STI transmission.

**C** – **C**ondom use: Correct and consistent use of male or female condoms during sexual intercourse to prevent STIs and unintended pregnancies.

**D** – **D**iagnosis (early detection, Avoid **D**rug): Early and regular screening for STIs, including asymptomatic infections, to enable timely treatment and reduce onward transmission.

**E** – **E**ducation: Comprehensive sexuality education to improve knowledge, attitudes, skills, and decision-making related to sexual and reproductive health.

## 3. RESULTS

The number of participants was 120 (Pre-test) and 54 (Post-test).

### 3.1. Sociodemographic Characteristics

At the pretest, the sample was slightly predominantly female (56.3%). Participants were health-related streamers, including physicians (13.8%), health technicians (12.1%), and biologists (11.5%). Participants were between the ages of 15 and 24 (47.7%), single (76.4%), without children (82.8%), and were Muslim (75.3%) or Christian (24.7%). Participants were from Mali (59.8%), followed by Benin (20.1%), Niger (11.5%) and Burkina Faso (8.6%) (**Table 2**).

**Table 2:** Distribution of Participants by Socio-demographic Characteristics.

| **Features** | **Modality** | **Frequency n=174** | **Percentage** |
| --- | --- | --- | --- |
| **Age** | 15-24 years old | 83 | 47,7 |
|  | ≥ 25 years old | 91 | 52,3 |
| **Gender** | Female | 98 | 56,3 |
|  | Male | 76 | 43,7 |
| **Nationality** | Mali | 104 | 59,8 |
|  | Benin | 35 | 20,1 |
|  | Niger | 20 | 11,5 |
|  | Burkina Faso | 15 | 08,6 |
| **Profession /**<br>**Stream of Study** | General Practitioners | 24 | 13,8 |
|  | Health Technicians | 21 | 12,1 |
|  | Biologists | 20 | 11,5 |
|  | Students (Nurse & Midwife) | 19 | 10,9 |
|  | High School Students | 18 | 10,3 |
|  | Students (MBA Public Health) | 14 | 08,0 |
|  | Students (Medicine) | 09 | 05,2 |
|  | Students (Pharmacy) | 09 | 05,2 |
|  | Midwives | 09 | 05,2 |
|  | Students (Electrical Engineering) | 05 | 02,9 |
|  | Journalists | 04 | 02,3 |
|  | Microbiologists | 04 | 02,3 |
|  | Epidemiologists | 04 | 02,3 |
|  | Chemists | 04 | 02,3 |
|  | Hygienists | 02 | 01,1 |
|  | Computer scientists | 02 | 01,1 |
|  | Pharmacists | 02 | 01,1 |
|  | Teachers | 02 | 01,1 |
|  | Building Engineer | 01 | 0,6 |
|  | Social Security Agent | 01 | 0,6 |
| **Marital Status** | Single | 133 | 76,4 |
|  | Married | 41 | 23,6 |
| **Paternity or**<br>**Maternity Status\*** | No | 144 | 82,8 |
|  | Yes | 30 | 17,2 |
| **Religion** | Islam | 131 | 75,3 |
|  | Christianity | 43 | 24,7 |

\*Paternity or maternity status = Having at least one child

### 3.2. General Knowledge of STIs and Genital Chlamydia

General knowledge about STIs improved slightly at post-test, although there was no significant difference (p = 0.63 and p = 0.82). Similarly, the proportion of participants able to name at least three modes of transmission of genital chlamydia increased from 41.7% to 50%, but did not reach statistical significance (p = 0.31 and p = 0.77). Symptom knowledge followed the same trend, increasing from 27.5% to 35.2% at post-test (p = 0.12 and p = 0.95) (**Table 3**).

**Table 3:** Distribution of Participants Based on their Knowledge on Genital Chlamydia.

| Item* | Category | Pre-test<br>n (%) | Post-test<br>n (%) | p-value |
| --- | --- | --- | --- | --- |
| **General Knowledge on STIs** | Very Good | 55 (47.0) | 27 (50.9) | 0.63 |
|  | Good | 58 (49.6) | 26 (49.1) | 0.82 |
|  | Poor | 04 (03.4) | 0 (0.0) | - |
| **Modes of Transmission** | At least 1 | 30 (25.0) | 12 (22.2) | 0.69 |
|  | At least 2 | 36 (30.0) | 15 (27.8) | 0.77 |
|  | At least 3 | 50 (41.7) | 27 (50.0) | 0.31 |
|  | At least 4 | 04 (03.3) | 0 (0.0) | - |
| **Symptoms** | None | 02 (01.7) | 0 (0.0) | - |
|  | At least 1 | 14 (11.7) | 08 (14.8) | 0.56 |
|  | At least 2 | 21 (17.5) | 04 (07.4) | 0.12 |
|  | At least 3 | 33 (27.5) | 19 (35.2) | 0.31 |
|  | At least 4 | 43 (35.8) | 19 (35.2) | 0.93 |
|  | At least 5 | 07 (05.8) | 04 (07.4) | 0.95 |
| **Complications**<br>**in Women** | At least 1 | 16 (13.3) | 09 (16.7) | 0.56 |
|  | At least 2 | 31 (25.8) | 08 (14.8) | 0.11 |
|  | At least 3 | 35 (29.2) | 15 (27.8) | 0.85 |
|  | At least 4 | 32 (26.7) | 20 (37.0) | 0.17 |
|  | At least 5 | 06 (05.0) | 02 (03.7) | 0.99 |
| **Complications**<br>**In Men** | At least 1 | 27 (22.5) | 20 (37.0) | **0.045\*** |
|  | At least 2 | 52 (43.3) | 11 (20.4) | **0.003\*\*** |
|  | At least 3 | 41 (34.2) | 23 (42.6) | 0.29 |
| **Complications**<br>**in Newborns** | None | 02 (01.7) | 0 (0.0) | - |
|  | At least 1 | 47 (39.2) | 16 (29.6) | 0.22 |
|  | At least 2 | 60 (50.0) | 31 (57.4) | 0.37 |
|  | At least 3 | 11 (09.2) | 07 (13.0) | 0.45 |
| **Prevention Methods** | At least 1 | 10 (08.3) | 02 (03.7) | 0.43 |
|  | At least 2 | 08 (06.7) | 03 (05.6) | 0.78 |
|  | At least 3 | 29 (24.2) | 05 (09.3) | **0.02\*** |
|  | At least 4 | 72 (60.0) | 42 (77.8) | **0.02\*** |
|  | At least 5 | 01 (0.8) | 02 (03.7) | 0.47 |

\*Modes of transmission: 1=Unprotected sexual intercourse, 2=Mother-to-child transmission 3=Direct contact with infected secretions 4= Other (specify) \*\*Symptoms: 1=Unusual vaginal discharge in women or urethral discharge in men, 2=Pain or burning when urinating, 3= Lower abdominal or pelvic pain, 4= Bleeding between periods or after intercourse \*\*\*Complications in women: 1=Pelvic inflammatory disease (PID); 2= Infertility, 3=Infertility, 4=Ectopic pregnancy (ECP), 5=Unintended pregnancies (UNDP), 6=Chronic pelvic pain \*\*\*\*Complications in men: 1=Epididymitis, 2=Infertility, 3=Infertility, 4=Sexual impotence\*\*\*\*\* Complications in newborns: 1= Severe conjunctivitis, 2= Neonatal pneumonia, 3= Umbilical hernia, 4=Other \*\*\*\*\* Prevention methods: 1=Condom, 2= Fidelity, 3= Regular screening, 4=Curative treatment with partner \*p < 0.05; \*\* p < 0.01.

### 3.3. Knowledge of Genital Chlamydia Complications

The proportions of those experiencing female complications also varied slightly between the two measurement times, with no significant change (p between 0.11 and 0.99). On the other hand, a significant improvement was observed for male complications: awareness of at least one complication increased (p = 0.045*) and knowledge of two major complications, such as sterility and infertility, increased significantly (p = 0.003**). Finally, knowledge of neonatal complications resulting from maternal infection increased slightly, although not significantly (p between 0.22 and 0.45) (**Table 3**).

### 3.4. Attitudes Towards Disclosure Genital Chlamydia to Partners and Stigma

Participants did not share information with partners about genital chlamydia, either before (89.2%) or after training (92.6%), while only a few discussed it (8.3% and 7.4%). The exchange between partners on STI status increased slightly, from 62.5% (Pretest) to 72.2% (Posttest) (p = 0.21). The proportion of participants who reported not stigmatizing people with STI increased slightly from 15.8% to 16.7% (p = 0.89) (**Table 4**).

**Table 4:** Distribution of Participants by Communication and Information Sharing with Sexual Partners.

| **Attitude / Behavior** | **Category** | **Pre-test n (%)** | **Post-test n (%)** | **p-value** |
| --- | --- | --- | --- | --- |
| **Sharing Information about Genital Chlamydia** | I don't know | 03 (02.5) | 0 (0.0) | – |
|  | No | 107 (89.2) | 50 (92.6) | 0.48 |
|  | Yes | 10 (08.3) | 04 (07.4) | – |
| **Discussion of STI Status between Partners** | No | 33 (27.5) | 11 (20.4) | 0.32 |
|  | Yes | 75 (62.5) | 39 (72.2) | 0.21 |
|  | Sometimes | 12 (10.0) | 04 (07.4) | – |
| **Stigmatization of People living with STIs** | I don't know | 05 (04.2) | 06 (11.1) | – |
|  | No | 19 (15.8) | 09 (16.7) | 0.89 |
|  | Yes | 96 (80.0) | 39 (72.2) | 0.26 |

### 3.5. Practices Related to STI Prevention and Care

Participants reported seeking immediate medical attention for STIs, both before (95%) and after training (96.3%) p=0.71. The use of health facilities for the management of STIs increased from 86.7% to 94.4% p=0.13, while the use of alternative solutions (traditional or similar) decreased from 13.3% to 5.6% p=0.13. The proportion of participants who had been tested for STIs remained stable (45% → 44.4%) p=0.95. Regular condom use improved slightly, with an increase in those who “always” reported using condoms (62.5% → 66.7%) p=0.69 and a decrease in those who “never” used them (19.2% → 16.7%) p=0.93. The ability to refuse unprotected sex increased from 63.3% to 68.5% p=0.51, and most participants reported having no multiple partners, both pretest (94.2%) and posttest (90.7%) (p=0.62) (**Table 5a**).

**Table 5a:** Distribution of Participants According to Prevention Practices and Use of STI Care.

| Attitude / Behavior | Category | Pre-test<br>n (%) | Post-test<br>n (%) | p-value |
| --- | --- | --- | --- | --- |
| **Use of a Health Professional in case of STIs** | Yes, immediately | 114 (95.0) | 52 (96.3) | 0.71 |
|  | Yes, but hesitantly | 06 (05.0) | 02 (03.7) | – |
| **Emergency Procedure in case of STI Symptoms** | Hospital or health center | 104 (86.7) | 51 (94.4) | 0.13 |
|  | Other / traditional / close care | 16 (13.3) | 03 (05.6) | 0.13 |
| **Ever Performed an STI Test** | No | 66 (55.0) | 30 (55.6) | 0.95 |
|  | Yes | 54 (45.0) | 24 (44.4) | 0.95 |
|  | Never | 23 (19.2) | 09 (16.7) | 0.69 |
| **Condom Use** | Sometimes | 15 (12.5) | 07 (13.0) | 0.93 |
|  | Rarely | 07 (05.8) | 02 (03.7) | 0.83 |
|  | Always | 75 (62.5) | 36 (66.7) | 0.60 |
| **Refusal to Have Sex without a Condom** | No | 21 (17.5) | 08 (14.8) | 0.66 |
|  | Yes | 76 (63.3) | 37 (68.5) | 0.51 |
|  | I don't know | 23 (19.2) | 09 (16.7) | 0.69 |
| **Having Multiple Sexual Partners** | No | 113 (94.2) | 49 (90.7) | 0.62 |
|  | Yes | 02 (01.7) | 02 (03.7) | – |
|  | Prefer not to answer | 05 (04.2) | 03 (05.6) | – |

### 3.6. Acceptability of STI Testing and Training Exposure

For the acceptability of on-demand STI testing, no “no” response was recorded at post-test. The proportion of participants who had ever been made aware of STIs increased slightly from 58.3% at the pre-test to 63% at the post-test. In contrast, the proportion of participants reporting having received STI training increased significantly, from 39.2% to 63% (p = 0.004) (**Table 5b**).

**Table 5b:** Distribution of Participants by STI Awareness and Previous Training.

| **Attitude / Behavior** | **Category** | **Pre-test n (%)** | **Post-test n (%)** | **p-value** |
| --- | --- | --- | --- | --- |
| **Acceptability of on-demand STI testing** | Yes | 85 (70.8) | 45 (83.3) | 0.08 |
|  | No | 02 (01.7) | 0 (0.0) | – |
|  | Maybe | 24 (20.0) | 07 (13.0) | – |
|  | I don't know | 09 (07.5) | 02 (03.7) | – |
| **Previous STI sensitization (including genital chlamydia)** | No | 50 (41.7) | 20 (37.0) | – |
|  | Yes | 70 (58.3) | 34 (63.0) | 0.56 |
| **Training on STIs (including genital chlamydia)** | No | 73 (60.8) | 20 (37.0) | 0.004** |
|  | Yes | 47 (39.2) | 34 (63.0) | – |

\*p < 0.05; \*\* p < 0.01

## 4. DISCUSSION

### 4.1. Characteristics of Participants and Summary of Key Findings

The study participants were predominantly female, young, single, and largely from health-related fields. Women accounted for 56.3% of the sample, most participants were single (76.4%), and only a minority reported having at least one child (17.2%). A substantial proportion had a health-related educational or professional background, which likely explains the relatively high baseline levels of knowledge and preventive practices observed before the intervention.

The educational workshops significantly increased participants’ formal training on STIs, including genital chlamydia, with the proportion reporting prior STI training rising from 39.2% to 63.0% (p = 0.004). Notable improvements were also observed in knowledge of male complications of genital chlamydia, particularly infertility and epididymitis, with statistically significant increases for awareness of at least one complication (p = 0.045) and at least two major complications (p = 0.003). In addition, participants’ ability to cite multiple prevention strategies based on the WHO ABCDE approach improved significantly (p = 0.02).

In contrast, care-seeking behaviors were already high at baseline and remained stable after the intervention, with timely prompt consultation of a health professional reported by more than 95% of participants at both time points. This ceiling effect likely limited the observable impact of the intervention on certain STI prevention practices. While this participant profile facilitates peer dissemination of acquired knowledge, it also limits the generalizability of our findings to all young people, particularly those who are out of school, less educated, or not connected to the health sector.

### 4.2. Inverted classroom and Small Group Facilitation

We intentionally provided the workshop booklet at least a week before the training and implemented our 5-day hybrid workshop using small group facilitation throughout. Prior research highlights six key factors that promote effective learning interactions: active participant engagement, participatory teaching approaches by trainers, collaborative teamwork within small groups, the integration of the local language, the hands-on nature of learning activities, and the availability of material resources. The combination of these elements fostered a collaborative and interdependent learning environment, supporting short-term enhancement of participants’ professional competencies (Dieudonné Musa Alokpo, 2023).

### 4.3. Hybrid Format of the Workshop

In resource-limited settings, internet connectivity can pose challenges for online training. However, online participation allows for a larger number of participants and can improve the cost-effectiveness of the intervention. Learners may have varying levels of comfort with simultaneous in-person and online formats, making preparatory sessions essential for online participants to ensure comparability in training outcomes (Emmanuelle Croze, 2019).

### 4.3. Knowledge of Genital Chlamydia

The workshops led to improvements in specific areas of knowledge related to genital chlamydia. Participants demonstrated better understanding of male complications and preventive methods, which are often underemphasized in STI education. However, general knowledge about STIs, modes of transmission, symptoms, and complications in women and newborns showed little change between pre- and post-test assessments. This limited variation is likely attributable to the relatively high baseline knowledge levels among participants, leaving little room for measurable improvement.

These findings suggest that short, intensive educational interventions may be particularly effective in addressing specific knowledge gaps rather than broadly improving already well-established concepts.

### 4.4. Attitudes, Partner Communication, and Prevention Practices

The intervention did not lead to significant changes in communication about genital chlamydia with sexual partners or in perceptions of STI-related stigma. Information sharing with partners remained low both before and after the workshops, and stigmatizing attitudes showed minimal change. These dimensions are deeply embedded in socio-cultural and religious norms, indicating that educational workshops alone are insufficient to produce substantial shifts in such attitudes (Lukumay et al., 2023).

Similarly, preventive practices and health service utilization showed little variation over time. Condom use, avoidance of multiple sexual partners, and timely consultation of health services were already relatively high at baseline. STI testing behavior remained unchanged, likely reflecting structural barriers such as limited access to screening services and the availability of diagnostic tools rather than deficits in knowledge.

### 4.5. Awareness, Training, and Comparison with the Literature

The marked increase in participants reporting having received STI training confirms the effectiveness of the workshops in strengthening perceived competence and formal knowledge. Although the improvement in acceptability of on-demand STI screening did not reach statistical significance, the upward trend is encouraging for prevention efforts.

These findings are consistent with existing literature showing that short-term educational interventions tend to improve targeted knowledge and training outcomes more effectively than complex behaviors or deeply rooted attitudes. The strong gains in knowledge related to male complications highlight the value of addressing often-neglected aspects of STI education. In contrast, changes in partner communication and stigma reduction typically require longer-term, repeated, and community-based interventions incorporating psychosocial components (Adohinzin et al., 2016; Guiella G, 2012).

### 4.6. Public Health Implications for Youth Sexual Health

From a public health perspective, the educational workshops contributed meaningfully to strengthening knowledge and training related to genital chlamydia among young people aged 15–24 years old. However, they are insufficient as standalone interventions to produce sustained changes in attitudes, communication, and sexual behaviors (Loyke Julie, 2023). For greater impact, such workshops should be integrated into comprehensive sexual health programs that include improved access to free or subsidized screening, community-level sensitization, long-term follow-up, and interventions targeting couples and families. Integrating genital chlamydia screening into premarital health check-ups could also be considered.

For young people’s sexual health, the workshops effectively enhanced awareness of key aspects of genital chlamydia, particularly male complications and prevention strategies. Better-informed youth are more likely to make safer sexual health decisions and act as peer educators within their communities (Silva et al., 2022). Nevertheless, the limited influence of intervention on partner communication, stigma, and testing behaviors underscores the need for sustained, multi-component approaches combining education, community engagement, and improved access to youth-friendly sexual health services to achieve long-term and sustainable reductions in STI transmission.

### 4.7. Study Limitations

This study has some limitations. Firstly, the absence of a control group restricts causal inference, and the short follow-up period did not allow assessment of long-term behavioral changes. Secondly, Reliance on self-reported data introduces potential social desirability bias. Finally, the sample composition - predominantly female, relatively well educated, and largely from the health sector - may have contributed to high baseline levels of knowledge and good practices, thereby limiting the generalizability of the findings to the broader population of young people, particularly those who are more vulnerable or less connected to health services.

## 5. CONCLUSION

This 5-day hybrid educational workshop effectively strengthened targeted knowledge on genital chlamydia and prevention strategies among young people aged 15–24 years old in Bamako, particularly regarding male complications and formal training on STIs. However, their short-term impact on attitudes, sexual communication, and certain preventive practices remained limited.

To enhance effectiveness and sustainability, future interventions should move beyond standalone workshops and be embedded within more comprehensive, longer-term sexual education programs. Such programs should (i) incorporate longitudinal follow-up, community- school-based and young couple-based approaches (ii) improve access - both in terms of availability and affordability - to genital chlamydia screening and youth-friendly sexual health services.

## Data Availability

All data produced in the present work are contained in the manuscript.

